# Characterizing the genetic architecture of drug response using gene-context interaction methods

**DOI:** 10.1101/2024.03.29.24305093

**Authors:** Michal Sadowski, Mike Thompson, Joel Mefford, Tanushree Haldar, Akinyemi Oni-Orisan, Richard Border, Ali Pazokitoroudi, Julien F. Ayroles, Sriram Sankararaman, Andy Dahl, Noah Zaitlen

## Abstract

Identifying the factors responsible for variability of treatment response is a central objective of clinical research. Specially designed pharmacogenomic studies have identified a handful of individual mutations modulating the effect of specific drugs. However, the extent to which drug response variability is driven by genetics is largely unknown, partly due to the small sample sizes of prospective pharmacogenomic trials. In this work, we develop a framework to study the genetic architecture of response to commonly prescribed drugs in large biobanks coupled with electronic health records. Our framework leverages concepts from gene-environment interaction testing, including novel interaction models at the level of genes, polygenic scores (PGS), and genome-wide heritability. We quantified the heritability of response to statins, metformin, warfarin, and methotrexate in 342,257 UK Biobank participants. Our results show that genetic variation modifies the primary effect of statins on LDL cholesterol (9% heritable) as well as its side effects on hemoglobin A1c and blood glucose (10% and 11% heritable, respectively). Next, we identified dozens of specific genes that modify drug response, which we then replicated in a retrospective pharmacogenomic study. Finally, we found that PGS accuracy varies up to 2-fold depending on treatment status, showing that the current approach of building PGS using mostly healthy individuals is likely to underperform in clinical contexts. Together, our results provide a framework for characterizing the genetic architecture of drug response using cross-sectional data.

## Main

Initiation of drug treatment poses a risk for adverse reactions and long term side effects, sometimes without guaranteed effectiveness for an individual patient^1–6^. Genetic testing holds promise for safer and more effective treatment by predicting each individual’s specific drug response^7–10^. To date, several large-effect pharmacogenomic genes have been identified^11–17^; these genes are commonly tested in the clinic to guide administration and dosing of certain medications^18–21^, which dramatically reduces the incidence of certain severe adverse drug reactions^15,17,22,23^.

More recently, genome-wide genetic data have been considered for clinical biomarkers of disease risk in the form of polygenic scores (PGS)^24–28^. PGS predict disease risk by aggregating many risk alleles identified by genome-wide association studies (GWAS)^29^. For some diseases, PGS have comparable performance to current clinical risk-prediction algorithms, at least in European ancestry individuals ^30–32^. However, genome-wide predictions for treatment response have not been developed, although they have been discussed extensively^33–36^.

Here, we build a framework to study genome-wide genetic effects on the primary and side effects of common drugs. Our approach leverages recent and novel methods for gene-environment interaction (GxE). Crucially, our methods apply to passively-obtained EHR data, enabling analyses of sample sizes far exceeding randomized controlled trials. We apply our approach to four of the most common drugs in the UK Biobank: statins, metformin, warfarin, and methotrexate. Our methods quantify genome-wide heritability of drug response, identify specific genes modifying drug response, and characterize the implications for clinical use of PGS. We replicate many of the gene-drug interactions in a longitudinal pharmacogenomic study of statins’ effects on LDL cholesterol^37,38^. Overall, our framework characterizes the genetic architecture of individual-level response to modifiable risk factors in passively-obtained EHRs.

## Results

### Study overview

We apply our framework to 342,257 unrelated white British individuals in the UK Biobank^39^ (Supplementary Material). We focus on four of the most commonly prescribed drugs in this dataset: statins, metformin, warfarin, and methotrexate. For each drug, we study phenotypes related to its primary effect as well as phenotypes related to its possible side effects (Table 1).

**Table 1:** Drug exposures and responses examined in this work.

| Drug exposure | Number of users (non-users) | Description | Primary effects | Side effects |
| --- | --- | --- | --- | --- |
| Statins | 56,169 (286,088) | Low-density lipoprotein (LDL) cholesterol lowering therapy prescribed to prevent atherosclerotic cardiovascular disease (ASCVD) events | LDL cholesterol, CVD<sup>72,73</sup> | Glucose, A1c, Type 2 diabetes (T2D)<sup>74–77</sup> |
| Metformin | 8,606 (333,651) | Antidiabetic therapy | Glucose, A1c<sup>78–81</sup> | Body mass index (BMI)<sup>82,83</sup>, LDL cholesterol, CVD<sup>84,85</sup> |
| Warfarin | 3,753 (338,504) | Anticoagulant therapy | Reticulocyte count, Hematocrit, Plateletcrit<sup>86–88</sup> | None examined |
| Methotrexate | 1,865 (340,392) | Antirheumatic therapy | C-reactive protein (CRP)<sup>89,90</sup> | None examined |

We first define and develop a novel approach to estimate the aggregate impact of common genetic variation on treatment response 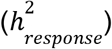. 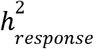 is the SNP heritability of the phenotype change after treatment (Fig. 1A, Methods). We estimate this parameter by post-processing results from GxEMM^40^, which was developed to estimate GxE-based heritability. GxEMM explicitly models treatment-dependent heteroscedasticity, which is essential for unbiased estimates of treatment-dependent heritability. Because this approach aggregates across the genome, it is powerful but does not identify specific genes.

**Fig. 1:**
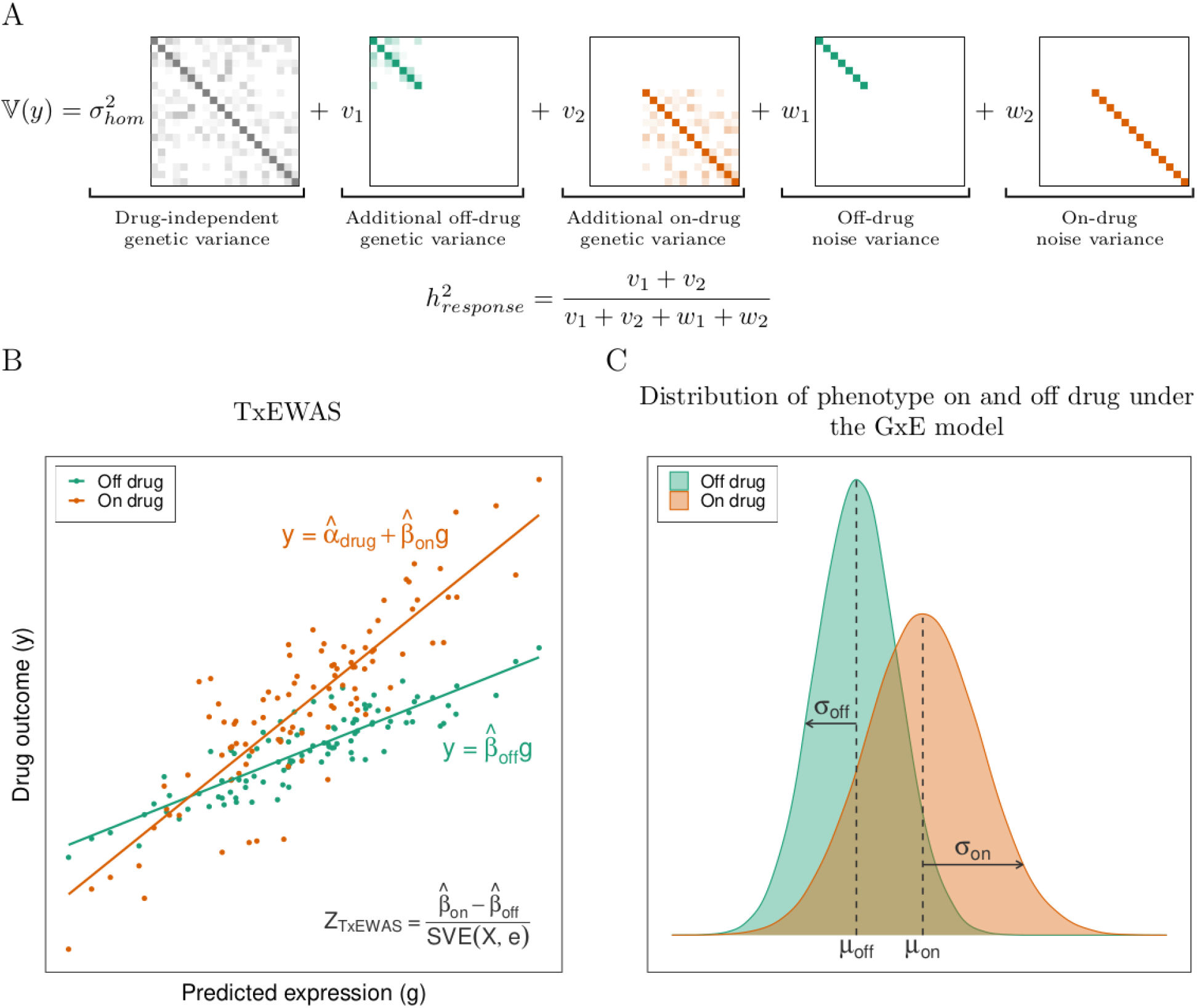
Schematic of the GxE framework to analyze treatment response in cross-sectional data. (A) We use GxEMM to estimate the heritability of treatment response 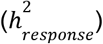 based on the genetic (*v*) and nongenetic (*w*) variances specific to treatment status (Methods). (B) We identify genes that modify treatment response using TxEWAS, a new method to estimate gene-level GxE interaction. TxEWAS genetically imputes gene expression and tests if this gene’s effect interacts with some “E”. (C) Statistical interactions with treatment status induce treatment-dependent heteroscedasticity that must be modeled in GxEMM and TxEWAS.

Next, we develop a new method called TxEWAS to identify specific genes that modify drug response. TxEWAS is a GxE extension of transcriptome-wide association studies (TWAS^41,42^). TxEWAS genetically imputes gene expression levels using reference transcriptomics data, as in TWAS, and then tests whether the imputed expression interacts with an environmental (“E”) variable (Fig. 1B, Methods). A major challenge in TxEWAS is accounting for treatment-dependent heteroscedasticity which we accomplish using the sandwich variance estimator (SVE)^43^ (Fig. 1C, Supplementary Fig. 1, Methods). Compared to SNP-level tests of GxE, TxEWAS improves power and interpretability.

Like all existing gene-environment interaction models, GxEMM and TxEWAS are susceptible to endogeneity bias because individuals’ treatments depend on their baseline phenotypes. We use theory, simulations, and additional data to characterize and account for this bias (Methods, Supplementary Material). Importantly, we test the statins-gene interaction effects on LDL in a retrospective longitudinal pharmacogenomic study, which validates these specific results as well as our cross-sectional approach.

Finally, we study the impact of treatment status, which varies significantly between individuals, on the performance of polygenic scores. We evaluate changes in PGS prediction accuracy by varying the proportion of treated individuals in the training and/or validation data.

### Primary effects and side effects of commonly-prescribed drugs are heritable

We estimated 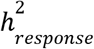 for statins’ effects on LDL cholesterol at 9% (Table 2). This is consistent with a prior estimate of 12% (SE=9%) derived by comparing first-degree relatives^37^. The heritability of statins’ effects on A1c and blood glucose were estimated at 10% and 11% respectively (Table 2). For comparison, the statin-independent heritabilities for these traits 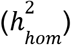 are 21%, 29% and 11% for LDL cholesterol, A1c and blood glucose, respectively (Table 2).

**Table 2:** Drug-independent heritability 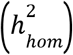, and heritability of drug response 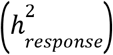 for a range of drug exposures and responses.

| Drug exposure | Response | $h^2_{hom}$ | $h^2_{response}$ | P value |
| --- | --- | --- | --- | --- |
| Statins | LDL cholesterol | 0.21 | 0.089 | $1.13 \times 10^{-30}$ |
| | A1c | 0.29 | 0.102 | $1.79 \times 10^{-6}$ |
| | Glucose | 0.11 | 0.111 | $2.26 \times 10^{-4}$ |
| Metformin | LDL cholesterol | 0.11 | 0.023 | 0.016 |
| | BMI | 0.28 | 0.170 | $2.51 \times 10^{-4}$ |

We next found that the 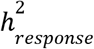 for metformin’s effects on LDL cholesterol and BMI are 2% and 17% respectively (Table 2), and the metformin-independent heritabilities are (11% and 28% respectively). However, we did not find significant heritability of response to warfarin or methotrexate, which was expected as we have lower power for less common drugs (Supplementary Table 1).

We next separately evaluated heritabilities in statin users vs non-users. We observe that the heritability of LDL cholesterol is much higher in individuals who do not take statins (41% vs 27%, Supplementary Table 2). This suggests that statins mitigate the genetic effects on LDL cholesterol present in untreated individuals (Supplementary Table 3). We found a qualitatively different pattern for the side effect of statins on A1c and blood glucose, where statin users had comparable or higher heritability. This suggests that statins activate or amplify genetic effects on blood sugar compared to untreated individuals. For metformin users, we found significantly higher heritability for BMI (51% vs 31%) and lower heritability for LDL cholesterol (11% vs 23%).

**Table 3:** Prediction accuracy of polygenic scores (PGS) trained in drug users, non-users, and a 50:50 mixture of both.

| Training | Prediction accuracy (Incremental $R^2$ [%]) | | | | | | | |
| --- | --- | --- | --- | --- | --- | --- | --- | --- |
|  | Statins |  |  |  | Metformin |  |  |  |
|  | A1c |  | LDL cholesterol |  | LDL cholesterol |  | BMI |  |
|  | On drug | Off drug | On drug | Off drug | On drug | Off drug | On drug | Off drug |
| On drug | 3.36<br>(0.23) | 2.32<br>(0.18) | 7.18<br>(0.30) | 12.60<br>(0.36) | 1.32<br>(0.37) | 3.24<br>(0.54) | 0.919<br>(0.286) | 0.0040<br>(0.0592) |
| Off drug | 2.56<br>(0.20) | 5.79<br>(0.28) | 7.98<br>(0.32) | 14.87<br>(0.39) | 2.75<br>(0.51) | 5.43<br>(0.67) | 0.041<br>(0.080) | 0.0077<br>(0.0636) |
| Agnostic | 2.60<br>(0.20) | 3.75<br>(0.23) | 5.86<br>(0.29) | 10.49<br>(0.36) | 2.25<br>(0.44) | 5.00<br>(0.66) | 0.255<br>(0.164) | 0.0931<br>(0.1078) |

### Identifying gene-drug interactions for primary and side effects

We next sought to identify specific genes that modify drug response. We found 156 genes that modify statins’ effects on LDL cholesterol (hFDR<10%, Fig. 2A, Supplementary Material). These genes include *PCSK9*, which is an LDL cholesterol-lowering drug target and has also been implicated in response to statin therapy^44,45^. Loss-of-function and gain-of-function variants in PCSK9 are known to reduce and elevate LDL cholesterol levels respectively^46,47^; statin therapy has been shown to increase serum PCSK9 levels, which can buffer statin LDL cholesterol-lowering effects. Consistent with these observations, we found that genetic effects that increase *PCSK9* expression reduce the effect of statins (Fig. 3A).

**Fig. 2:**
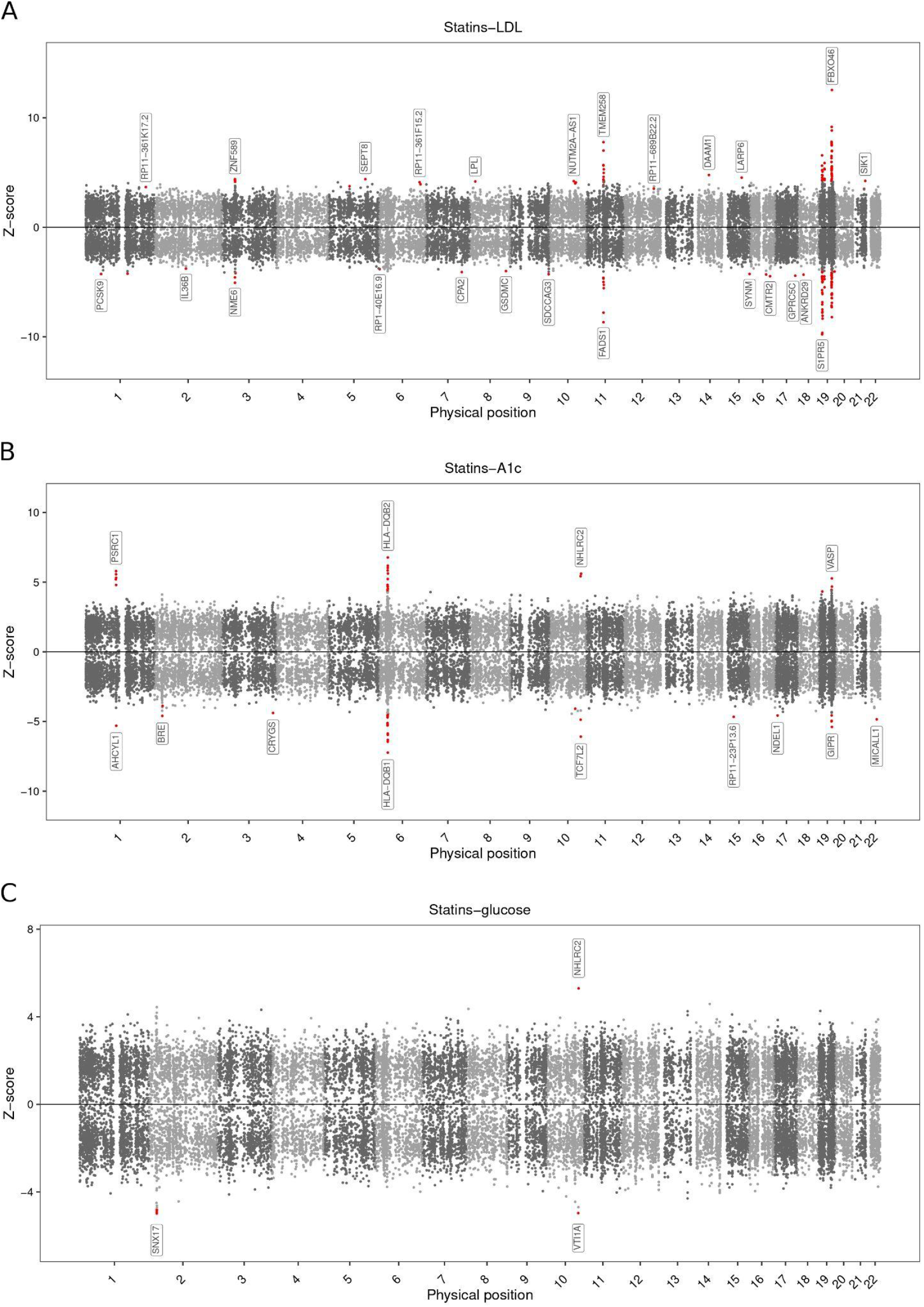
Manhattan plots of gene-statin interactions for low-density lipoprotein (LDL) cholesterol (primary effect), and hemoglobin A1c and blood glucose (side effects). (A) Manhattan plot of gene-statin interaction effects for LDL cholesterol. Each point represents a single gene, with physical position plotted on the x axis and standardized effect size plotted on the y axis. The most extreme effect across tissues is shown for each gene. Significant associations are highlighted in red, and the strongest associations on each chromosome are labeled. (B) and (C) follow (A), but for A1c and blood glucose.

**Fig. 3:**
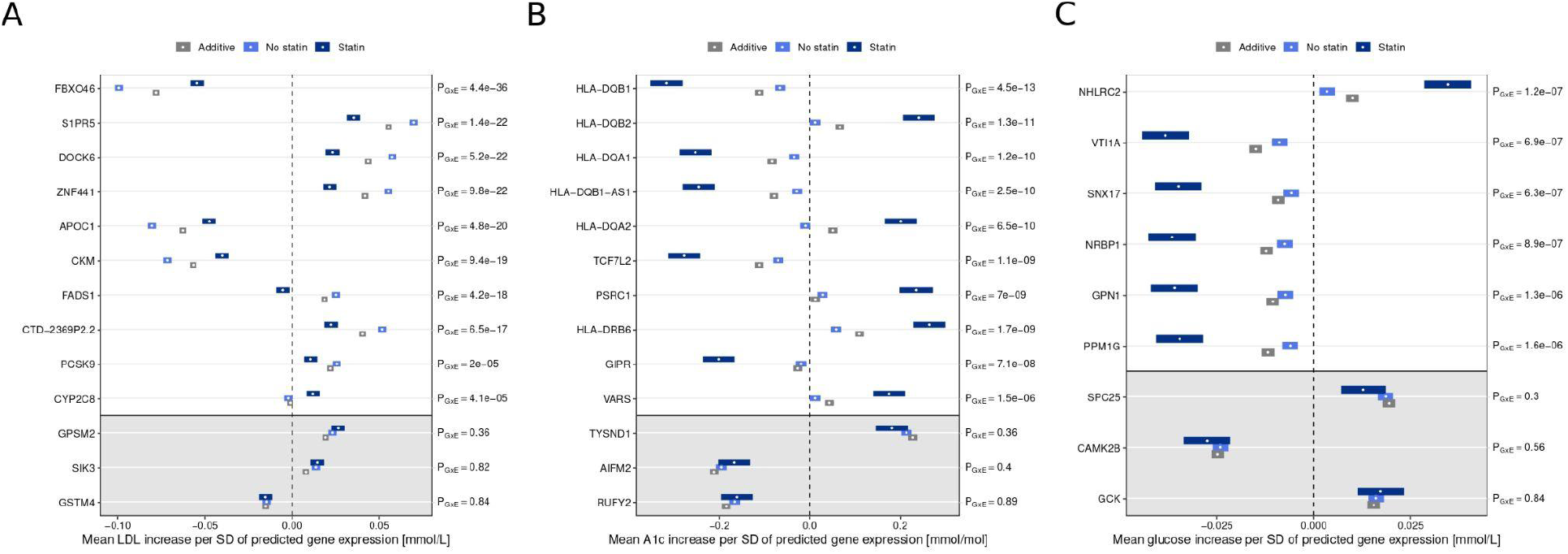
Gene-statin interaction effect sizes for low-density lipoprotein (LDL) cholesterol, hemoglobin A1c, and blood glucose. (A) Estimated effect sizes of selected genes on LDL cholesterol in statin users, non-users, and from a standard additive model. The top 10 genes have significant interaction effects; for comparison, the bottom three genes are only additively significant. (B) and (C) are similar to (A), but for A1c and blood glucose.

Of the 156 genes significantly interacting with statins, 131 also have significant additive effects (hFDR<10%). Interestingly, all 131 interaction effects have opposite signs to the main effects (Fig. 3A and Supplementary Fig. 2). That is, statins uniformly buffer these genetic effects on LDL cholesterol. This is consistent with our observation that LDL cholesterol has higher heritability in statin non-users than users (Supplementary Table 2).

We next tested which genes modify the side effects of statins on A1c and blood glucose, and we identified 53 and 6, respectively (Fig. 2B and 2C). 28 of the 53 genes with statin-dependent effects on A1c lie in the highly complex MHC region, and it is likely that many of these are not causal^48^. One example gene outside the MHC is *GIPR*, which is known to regulate insulin levels in the presence of elevated glucose in mice^49^ and is associated with increased risk of hyperinsulinemia after an antipsychotic treatment^50^. All six genes that modify statins’ effects on glucose overlap the statin-dependent A1c loci (Fig. 2C).

Of the 53 genes with statin-dependent effects on A1c, 36 have significant additive effects. However, unlike the genes modifying statins’ effects on LDL cholesterol, only four of them have an interaction effect with opposite sign to the primary effect (Fig. 3B and Supplementary Fig. 3). Similarly, only three of the six genes with statin-dependent effects on glucose exhibit significant additive effects, and all have the same sign as the corresponding interaction effects (Fig. 3C). Broadly, this suggests that some genetic effects on A1c and glucose are amplified by statins, while others are dampened.

We next analyzed metformin, warfarin, and methotrexate for gene-drug interactions. Although these have many fewer users (Table 1), we identified three gene-warfarin interactions effects on reticulocyte count (*HIF3A*, implicated in the response to hypoxia^51^; *ITGA1*, shown to be upregulated at high oxygen levels in the environment^52^; and *AL049542*.*1*) and one gene-methotrexate interaction effect on C-reactive protein levels (*C6orf164*).

The results are highly concordant if we exclude individuals who take combinations of the aforementioned drugs (Supplementary Table 4, Supplementary Material).

### Replicating gene-drug interactions in a pharmacogenomic study

The UK Biobank EHR data is passively obtained from an observational cohort and may suffer from confounding due to endogeneity in treatment status–sick individuals are more likely to be on treatments, and this may be driven in part by genetics. Therefore, we validated our approach by replicating the gene-level interactions for statins’ primary effect in a pharmacogenomic study^37,38^ (Methods). Of the 156 significant genes that we identified from cross-sectional data, 155 could be studied in the replication cohort (Supplementary Material). We found that 36/155 genes replicated (hFDR<10%), and that the remaining genes were significantly enriched for low P values<0.1 (binomial test p=0.002).

### Gene-drug interactions impact polygenic prediction accuracy

Gene-drug interactions violate underlying assumptions of PGS because they assume that genetic effects are independent of treatment status. We assessed the impact of this bias on PGS performance by varying the proportion of individuals on a drug in the training and/or testing cohorts, keeping the training sample size fixed. We focused on statins and metformin because they had treatment response heritability (Supplementary Table 5 and Methods).

First, we evaluated PGS for A1c as a function of statin use (Table 3). We find that statin users are better predicted by PGS trained on statin users, and vice versa for statin non-users.

Concretely, prediction accuracy of PGS for A1c in statin users increases by 31% when it is trained in treated vs untreated individuals. While this is intuitive, it need not hold in general. For example, if two groups share identical genetic effects but have different levels of non-genetic noise, the PGS should always be trained in the less-noisy group. Indeed, we observe this pattern for LDL cholesterol, which has higher heritability in statin non-uses than users, and we find that training PGS in non-users is optimal for predicting in users (Table 3). We performed extensive simulations to confirm these results (Supplementary Table 6, Supplementary Material). Finally, we evaluated an “agnostic” PGS built from a mix of users and non-users without accounting for statins, and we found that this PGS performed worst of all (Table 3). This illustrates an unappreciated limitation of standard approaches to building PGS in biobanks.

We found qualitatively similar results for PGS dependence on metformin, though they had smaller sample size and were weaker: BMI for users were better predicted using PGS built from users, while LDL cholesterol was always better predicted using non-users (Table 3). Overall, PGS blind to drug use will have heterogeneous accuracies between treated and untreated patients.

## Discussion

We quantified the genome-wide contribution of genetic variation to drug response 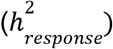 for four of the most commonly prescribed drugs worldwide. We identified specific genes driving this variation, and we validated the gene-statin interaction effects on LDL cholesterol in a longitudinal pharmacogenomic study. We found that such genetic effects on drug response have downstream implications for PGS, which are moving toward clinical use. In particular, we showed that current PGS will often underperform in the clinic because they are biased toward untreated individuals. While our paper focuses on drug treatments, we note that our novel framework can characterize the genetic basis of any covariate’s effect, including sex/gender, age, or modifiable risk factors.

Our results suggest that genome-wide genetic variation broadly modifies drug response. This is an important extension of pharmacogenomic studies, which usually focus on large-effect genes directly involved in drug metabolism. This extension is consistent with the overall arc of human genetic studies, where first large-effect genes are identified and then, as sample sizes grow and methods mature, genome-wide signals are identified^53,54^. Importantly, our results identify an unappreciated limitation of the clinical use of PGS, which are systematically less predictive in treated individuals than healthy controls. This complements other known limitations of PGS, including limited transferability across socio-economic status, age, sex^55,56^, and ancestry^57,58^, which could also be driven in part by gene-environment interactions^59^. On the other hand, our results pave a path to developing context-aware PGS^60^, which could directly predict an individual’s response to common drugs.

Our study has several limitations. First, it has focused on the UK Biobank, which is a cross-sectional cohort with non-random allocation of drugs. This raises the possibility of endogeneity biases causing false positives or false negatives, where our results reflect causes of drug prescription rather than its consequences. Nonetheless, we have validated many of our results in a longitudinal pharmacogenomic study, which took steps to reduce biases from dosing and baseline LDL cholesterol levels. More importantly, for many traits, the results are inconsistent with simple endogeneity-driven biases because we observed both positive and negative interaction effects. A related limitation is that our cross-sectional approach to estimate heritability only provides a lower bound to 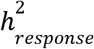 (Supplementary Material). In the future, methods to formally account for such endogeneity would give more precise estimates of treatment response heritability. Second, statistical genetic interactions need not reflect biochemical interactions between a gene and drug. This contrasts with large-effect pharmacogenomic genes, which typically encode enzymes that directly metabolize the drug. Third, the TxEWAS method is liable to detect genes or tissues that are merely correlated with causal genes of tissues. In the future, established TWAS methods to fine-map causal genes^48^ or tissues^61–63^ could be adapted to the TxEWAS setting. Fourth, due to biases in available data, we only analyzed individuals of European ancestry; more ancestrally diverse data are needed to obtain more generally applicable results.

In conclusion, we provided evidence for substantial polygenic contributions to drug response, and showed how large-scale cross-sectional studies like the UK Biobank can be used to estimate genetic effects on drug response. Although validation with randomized controlled trials is needed before drawing definitive conclusions about causal genetic effects on treatment response, our results demonstrate that cross-sectional data can generate compelling hypotheses on genetic modifiers and statistical predictions for treatment response. It is important, as advances in pharmacogenomics cannot be made with sole input from randomized controlled trials^64^, which have strict inclusion and exclusion criteria, cannot always be performed due to ethical issues, and are limited to small sample sizes due to high cost. Based on these results, we envision that novel PGS approaches incorporating treatment information will provide actionable clinical guidelines for optimizing primary effects and minimizing harmful side effects of drugs.

## Methods

### UK Biobank Data

Analyses presented in this work were performed in the UK Biobank population of 342,257 unrelated white British individuals (Supplementary Material).

For heritability and PGS analyses, we used 579,566 UK Biobank variants with minor allele frequency (MAF) larger than 0.01, Hardy-Weinberg equilibrium (HWE) test P value below 10^−10^, and imputation INFO score of 1. For the TxEWAS analysis, UK Biobank SNPs that matched eQTLs trained in the GTEx consortium were used.

UK Biobank fields used to retrieve phenotypes can be found in the Supplementary Material. For non-binary outcomes, we discarded measurements greater than five standard deviations from the mean, with the assumption that such extreme levels were results of non-modeled circumstances. The only exception was C-reactive protein levels, which were inverse normally transformed. For disease outcomes, we only retained diagnoses recorded after the date of the initial assessment with the UK Biobank initiative (when the information about medication use was collected).

The main analyses reported in this work were performed using the following covariates: age, sex, birth date, Townsend deprivation index, and the first 16 genetic PCs^57^. We additionally accounted for the measuring device type when an outcome required it, which was the case for hematocrit, plateletcrit, and reticulocyte count. All non-binary covariates were standardized (transformed to mean-zero, variance 1) before calculating interaction variables.

### Quantifying the heritability of treatment response

GxEMM quantifies the heritability contributed by genome-wide additive effects and genome-wide GxE effects. The general GxEMM model for phenotype *y*_*i*_ in environment *k* (i.e.*z*_*i*_ = *k*) is:

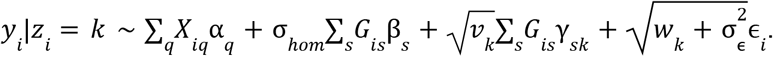

In this model, *X* are covariates with fixed effects α, and *G* is the genotype matrix, with additive effects β. We assume that β and the noise, ϵ, are i.i.d. standard normal, and the additive heritability is determined by the genetic and noise variances, 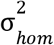 and 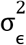. GxEMM additionally captures SNP-environment interaction effects, γ, which are also assumed i.i.d. standard normal.

Further, GxEMM allows environment-specific genetic (*v*_*k*_) and noise (*w*_*k*_) variances. If the phenotype is scaled to variance 1, 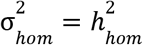.

Here, we use treatment status as the “environment” in order to quantify the heritability due to treatment-specific effects. We approximate the heritability of treatment response, Δ*y*_*i*_ = *y*_*i*_ (*z*_*i*_ = 1) − *y*_*i*_ (*z*_*i*_ = 0), by:

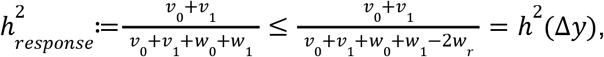

where *w*_*r*_ captures the covariance in effect sizes for unmodeled risk factors between treated/untreated states (Supplementary Material).

For warfarin, methotrexate, and metformin, we studied a sample of 30,000 individuals that included all users of that drug and an accordingly-sized random draw of non-users. To assess stability of our results, we repeated the analysis five times by randomly resampling non-users, and reported results from the sample with median additive heritability 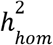, Supplementary Fig. 4). Because statins are much more common, we instead randomly split all 342,257 individuals into 11 non-overlapping subsets and meta-analyzed the results. This is a common approach employed in biobank-scale datasets to reduce computational complexity^65^.

### Identifying genes responsible for variable drug response

TxEWAS extends transcriptome-wide association studies (TWAS^41,42^) to test gene-environment interactions. The TxEWAS framework involves two major steps: First, gene expression levels of each gene are genetically imputed using a reference dataset. Second, the interaction effect, γ, between imputed gene expression and the drug is tested in the regression model:

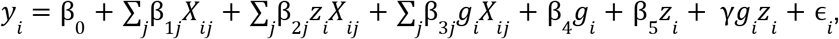

where ϵ_*i*_ ∼ *N*(0, σ^2^); *z*_*i*_ and *g*_*i*_ are the drug use indicator and imputed expression of some gene for individual *i*, respectively, and *X* is a matrix of covariates.

For binary phenotypes, the interaction effect is tested in the logistic regression model with the same covariates (Supplementary Material). In both models, the variance of the effect size estimates is estimated with the robust sandwich variance estimator to control for heteroskedasticity and/or misspecification of the functional form of the environmental factor^66,67^ (Supplementary Material).

In this study, we imputed gene expression into the UK Biobank using the 48 tissues from the GTEx consortium, and we used hierarchical FDR^68–70^ (hFDR<10%) to account for multiple hypothesis testing across genes and tissues (Supplementary Material).

### Replication in a pharmacogenomic study

We initially discovered gene-drug associations in cross-sectional data using TxEWAS. To validate these discoveries, we performed ordinary TWAS on the change in LDL cholesterol after statin initiation in an external pharmacogenomic study (we term this analysis PGx TWAS). More concretely, we used data from a longitudinal study of 28,616 individuals with European ancestries from the Kaiser Permanente GERA cohort (Genetic Epidemiology Research on Adult Health and Aging)^37,38^. For every TxEWAS interaction gene identified in the UK Biobank cohort, we calculated the PGx TWAS statistic in all available GTEx tissues, and employed an hFDR correction to call statistically significant genes at FDR<10% (Supplementary Material).

### Assessing implications for polygenic scores in clinical practice

PGS are weighted sums of risk alleles optimized to predict some training dataset. This makes PGS depend on characteristics of the training data, such as ancestry^58^, age^55^, or sex^56^. We assessed the accuracy of PGS as a function of drug use, including PGS trained in users and tested in non-users and vice-versa. In the main analysis, we varied the proportion of individuals on a drug in the training cohort, keeping the sample size fixed. We evaluated additional scenarios in the Supplementary Material. We fit PGS using a fast implementation of penalized linear regression with the lasso penalty^57,71^ and we measured prediction accuracy by the incremental *R*^2^ over baseline covariates. Standard errors around the estimates were calculated using bootstrap.

## Data Availability

No data was produced in the present work. This research was conducted using the UK Biobank Resource under application 33127. We thank the participants of the UK Biobank for making this work possible.

https://github.com/michalsad/txewas_scripts

https://github.com/andywdahl/gxemm

