## Supplementary Material for "Characterizing the genetic architecture of drug response using gene-context interaction methods"

### 1 Figures

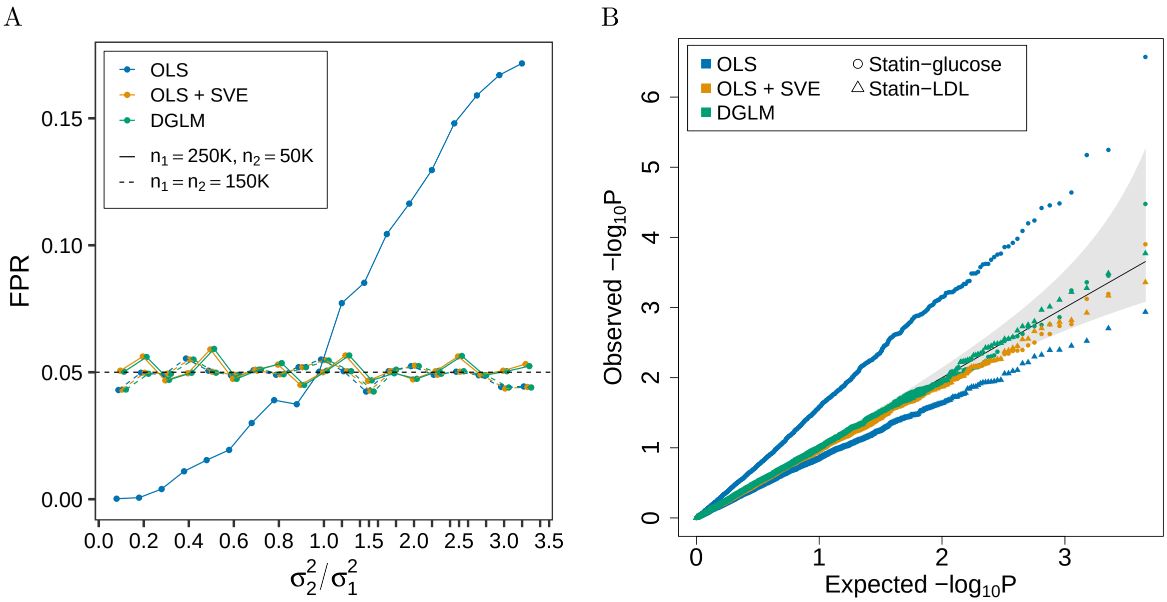

Supplementary Fig. 1: Evaluation of the type I error rate for the GxE effect estimated with the OLS model, the OLS model using robust standard errors (OLS + SVE) and the DGLM. (A) False positive rate (FPR) of GxE as a function of the ratio between phenotype variances in two environments: unexposed (of size $n_1$ and phenotype variance $\sigma_1^2$), and exposed (of size $n_2$ and phenotype variance $\sigma_2^2$). The nominal FPR of 5% is marked by the black dashed line. (B) Quantile-quantile plot comparing the null expected P values (x-axis) to the observed GxE P values after permuting real data from the UK Biobank (y-axis). The permutation permutes imputed expression of 4,516 genes and then tests their interaction with statins on blood glucose (circles) or LDL cholesterol (triangles).

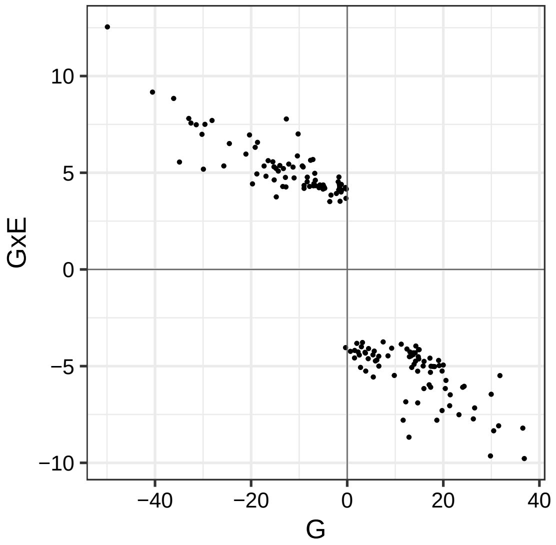

Supplementary Fig. 2: Z-scores for the main (G) and interaction (GxE) effects of genes whose interactions with statins were significantly associated with LDL cholesterol in TxEWAS. For each gene, we plot the estimates corresponding to the tissue with the strongest interaction p-value.

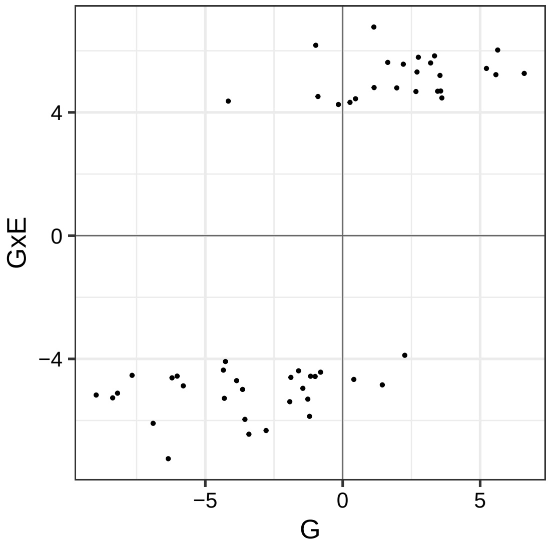

Supplementary Fig. 3: Z-scores for the main (G) and interaction (GxE) effects of genes whose interactions with statins were significantly associated with A1c in TxEWAS. For each gene, we plot the estimates corresponding to the tissue with the strongest interaction p-value.

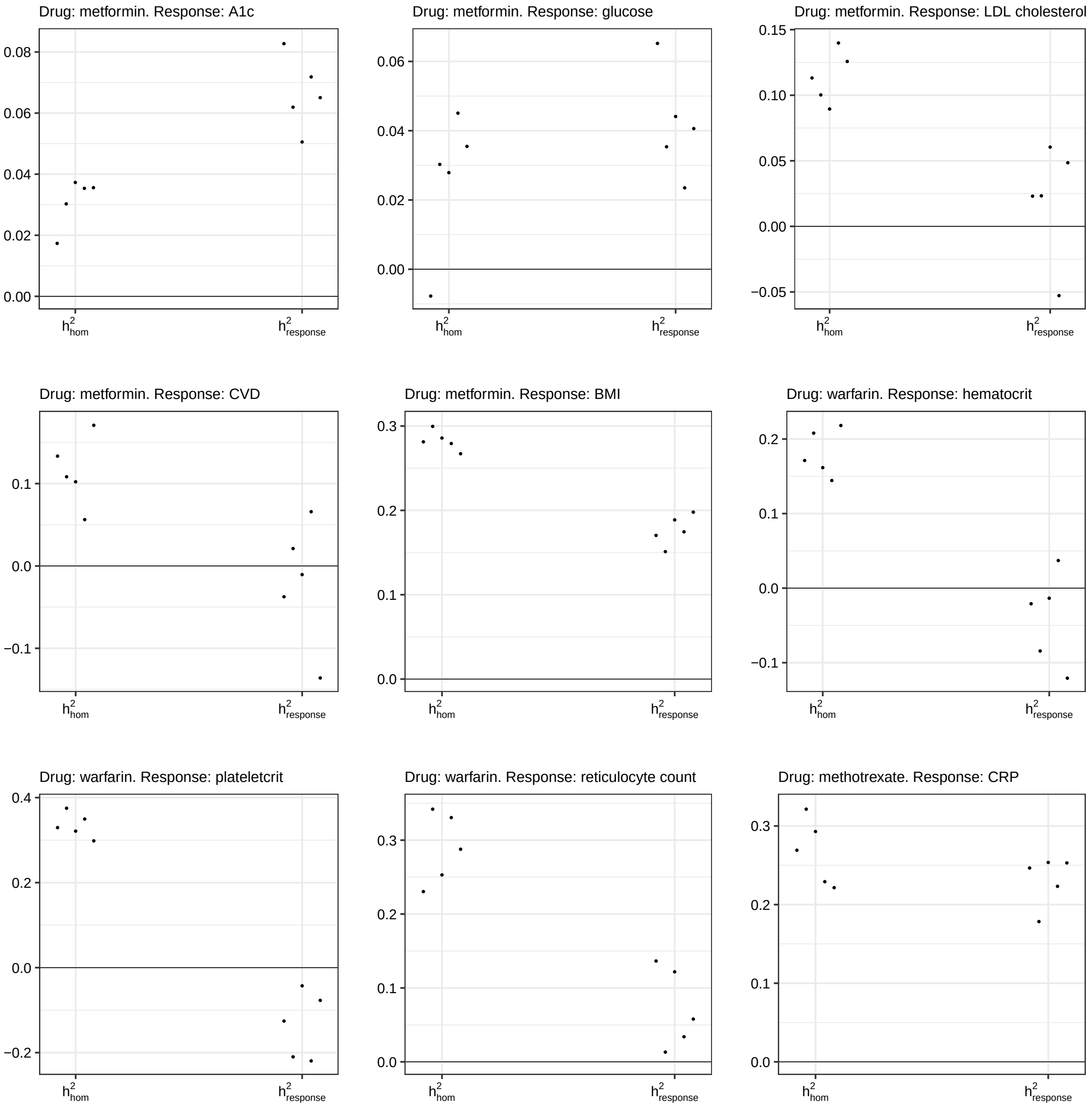

Supplementary Fig. 4: Estimation of drug-independent heritability $\left(h^2_{hom}\right)$, and heritability of drug response $\left(h^2_{response}\right)$ repeated five times with randomly resampled non-users.

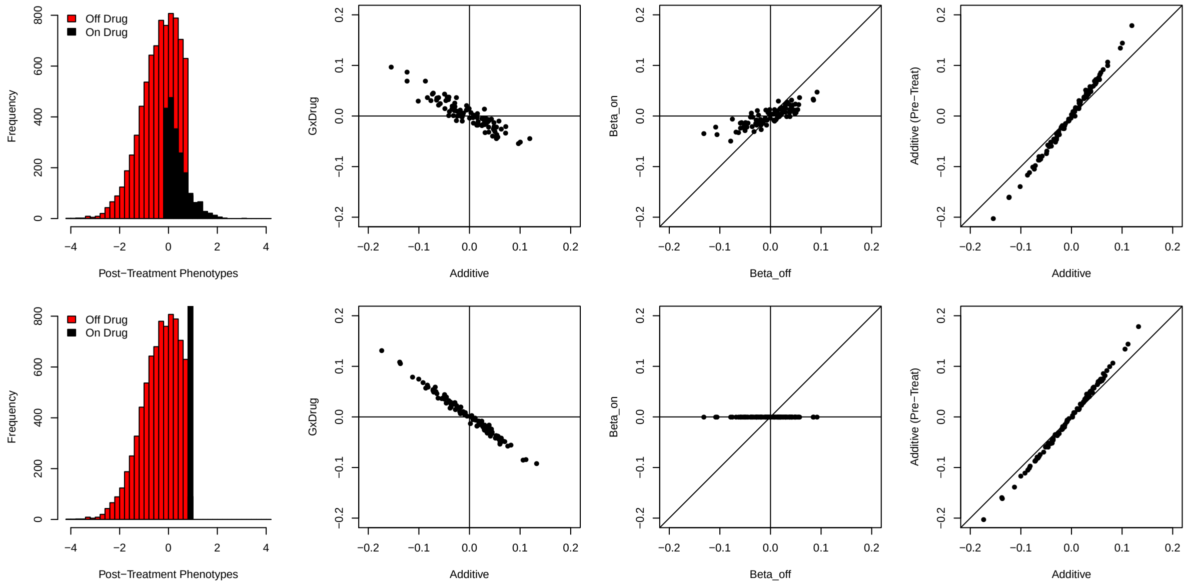

Supplementary Fig. 5: Endogeneity bias simulations. Top row: Simulation assumes that treatments have an equal additive effect on all individuals. Bottom row: Simulation assumes that treatments return all individuals to the treatment threshold, regardless their initial phenotypes. First column: Cross-sectional phenotype distributtion, stratified by treatment status. Second column: Comparison of estimated additive vs interaction effect sizes. Third column: Comparison of estimated effect sizes in treated vs untreated individuals. Fourth column: comparison of additive effect estimated on pre-treatment phenotypes vs cross-sectionally observed phenotypes containing a mix of treated and untreated individuals.

#### 2 Tables

Supplementary Table 1: Drug-independent heritability $\left(h^2_{hom}\right)$, and heritability of drug response $\left(h^2_{response}\right)$ for a range of drug exposures and responses.

| Drug exposure | Response | $h^2_{hom}$ | $h^2_{response}$ | P value |
|---|---|---|---|---|
| Statins | LDL cholesterol | 0.208 | 0.0892 | $1.13 \times 10^{-30}$ |
| | CVD | 0.086 | 0.0118 | 0.797 |
| | A1c | 0.285 | 0.1018 | $1.79 \times 10^{-6}$ |
| | Glucose | 0.112 | 0.1114 | $2.26 \times 10^{-4}$ |
| | T2D | 0.319 | -0.0119 | 0.936 |
| Metformin | A1c | 0.035 | 0.0719 | 0.516 |
| | Glucose | 0.030 | 0.0354 | 0.907 |
| | LDL cholesterol | 0.113 | 0.0230 | 0.016 |
| | CVD | 0.108 | 0.0211 | 0.924 |
| | BMI | 0.281 | 0.1703 | $2.51 \times 10^{-4}$ |
| Warfarin | Hematocrit | 0.171 | -0.0209 | 0.991 |
| | Plateletcrit | 0.329 | -0.1257 | 0.291 |
| | Reticulocyte count | 0.288 | 0.0579 | 0.627 |
| Methotrexate | CRP | 0.269 | 0.2466 | 0.337 |

Supplementary Table 2: Off-drug $\left(h^2_{\text{off}}\right)$ and on-drug $\left(h^2_{\text{on}}\right)$ heritability estimates for a range of drug exposures and responses.

| Drug exposure | Response | $h^2_{\text{off}}$ (SE) | $h^2_{\text{on}}$ (SE) |
|---|---|---|---|
| Statins | LDL cholesterol | 0.4125 (0.0080) | 0.270 (0.034) |
| | CVD | 0.1029 (0.0739) | 0.075 (0.081) |
| | A1c | 0.3170 (0.0075) | 0.286 (0.034) |
| | Glucose | 0.0924 (0.0068) | 0.213 (0.033) |
| | T2D | 0.2820 (0.4547) | 0.119 (0.035) |
| Metformin | A1c | 0.3040 (0.0568) | 0.081 (0.147) |
| | Glucose | 0.0758 (0.0302) | 0.057 (0.106) |
| | LDL cholesterol | 0.2294 (0.0317) | 0.114 (0.075) |
| | CVD | 0.1975 (0.3253) | 0.067 (0.137) |
| | BMI | 0.3073 (0.0300) | 0.513 (0.073) |
| Warfarin | Hematocrit | 0.2647 (0.0296) | 0.153 (0.187) |
| | Plateletcrit | 0.3936 (0.0231) | 0.180 (0.112) |
| | Reticulocyte count | 0.3385 (0.0209) | 0.337 (0.138) |
| Methotrexate | CRP | 0.3457 (0.0202) | 0.549 (0.248) |

Supplementary Table 3: Variance decomposition with GxEMM for a range of drug exposures and responses.

| Drug exposure | Response | $\sigma^2_{\text{g(hom)}}$ (SE) | $\sigma^2_{\text{g(off)}}$ (SE) | $\sigma^2_{\text{g(on)}}$ (SE) | $\sigma^2_{\text{e(off)}}$ (SE) | $\sigma^2_{\text{e(on)}}$ (SE) |
|---|---|---|---|---|---|---|
| Statins | LDL cholesterol | 0.2077 (0.0090) | 0.1414 (0.0110) | -0.0483 (0.0216) | 0.4973 (0.0068) | 0.4528 (0.0214) |
| | CVD | 0.0860 (0.0400) | -0.0049 (0.0501) | 0.0315 (0.1101) | 0.7637 (0.2682) | 1.4621 (0.4331) |
| | A1c | 0.2849 (0.0132) | -0.0840 (0.0138) | 0.2989 (0.0698) | 0.4328 (0.0049) | 1.4629 (0.0698) |
| | Glucose | 0.1116 (0.0131) | -0.0450 (0.0137) | 0.3361 (0.0695) | 0.6600 (0.0052) | 1.6615 (0.0694) |
| | T2D | 0.3187 (0.0481) | -0.0825 (0.0647) | 0.0473 (0.0983) | 0.2676 (0.7135) | 2.7246 (0.3624) |
| Metformin | A1c | 0.0354 (0.0328) | 0.0398 (0.0352) | 0.0394 (0.1348) | 0.1720 (0.0144) | 0.8505 (0.1368) |
| | Glucose | 0.0303 (0.0334) | -0.0054 (0.0344) | 0.0886 (0.2220) | 0.3037 (0.0113) | 1.9657 (0.2222) |
| | LDL cholesterol | 0.1132 (0.0233) | 0.0778 (0.0337) | -0.0513 (0.0436) | 0.6415 (0.0264) | 0.4803 (0.0412) |
| | CVD | 0.1083 (0.0929) | 0.0518 (0.1310) | -0.0072 (0.1826) | 0.6507 (1.1650) | 1.4190 (1.2552) |
| | BMI | 0.2813 (0.0292) | -0.0618 (0.0350) | 0.2706 (0.0828) | 0.4948 (0.0216) | 0.5222 (0.0782) |
| Warfarin | Hematocrit | 0.1712 (0.0427) | -0.0023 (0.0459) | -0.0241 (0.1808) | 0.4694 (0.0190) | 0.8145 (0.1807) |
| | Plateletcrit | 0.3295 (0.0369) | 0.0108 (0.0409) | -0.1570 (0.1009) | 0.5243 (0.0199) | 0.7850 (0.1082) |
| | Reticulocyte count | 0.2877 (0.0372) | 0.0400 (0.0417) | 0.0389 (0.1369) | 0.6404 (0.0203) | 0.6433 (0.1338) |
| Methotrexate | CRP | 0.2692 (0.0536) | 0.0550 (0.0563) | 0.2986 (0.2573) | 0.6135 (0.0189) | 0.4670 (0.2559) |

Supplementary Table 5: Prediction accuracy of PGS trained in drug users and non-users.

| PGS | Prediction accuracy (Incremental $R^2$ [%]) | | | | | | | |
|---|---|---|---|---|---|---|---|---|
| | Statins | | | | Metformin | | | |
| | A1c | | LDL cholesterol | | LDL cholesterol | | BMI | |
| | On drug | Off drug | On drug | Off drug | On drug | Off drug | On drug | Off drug |
| on-drug-PGS | 3.36 (0.23) | 2.32 (0.18) | 7.18 (0.30) | 12.60 (0.36) | 1.32 (0.37) | 3.24 (0.54) | 0.919 (0.286) | 0.0040 (0.0592) |
| off-drug-PGS | 2.56 (0.20) | 5.79 (0.28) | 7.98 (0.32) | 14.87 (0.39) | 2.75 (0.51) | 5.43 (0.67) | 0.041 (0.080) | 0.0077 (0.0636) |
| standard-PGS | 2.60 (0.20) | 3.75 (0.23) | 5.86 (0.29) | 10.49 (0.36) | 2.25 (0.44) | 5.00 (0.66) | 0.255 (0.164) | 0.0931 (0.1078) |
| half-PGS | 2.80 (0.21) | 3.75 (0.22) | 7.67 (0.32) | 13.59 (0.38) | 2.14 (0.45) | 5.24 (0.67) | 0.193 (0.141) | -0.0205 (0.0370) |
| all-PGS | 4.70 (0.26) | 5.62 (0.25) | 8.62 (0.32) | 15.66 (0.42) | 3.01 (0.53) | 6.63 (0.74) | 0.212 (0.148) | 0.0477 (0.0843) |
| on-drug-PGS + off-drug-PGS | 4.12 (0.24) | 6.02 (0.28) | 8.15 (0.34) | 14.93 (0.40) | 2.74 (0.51) | 5.42 (0.66) | 0.959 (0.299) | 0.0120 (0.0851) |
| all-PGS + on-drug-PGS + off-drug-PGS | 4.72 (0.26) | 6.82 (0.30) | 8.65 (0.33) | 15.82 (0.40) | 3.12 (0.55) | 6.70 (0.79) | 0.950 (0.305) | 0.0341 (0.1029) |

Supplementary Table 6: Simulation of polygenic prediction in individuals on and off a treatment.

| PGS | Prediction accuracy $(R^2)$ | | | |
|---|---|---|---|---|
| | Scenario 1 | | Scenario 2 | |
| | On drug | Off drug | On drug | Off drug |
| on-drug-PGS | 0.0881 | 0.2654 | 0.3613 | 0.2091 |
| off-drug-PGS | 0.0940 | 0.2838 | 0.2548 | 0.2955 |
| all-PGS | 0.0942 | 0.2844 | 0.3379 | 0.2699 |

### 3 Variance decomposition analysis with GxEMM

#### 3.1 The GxEMM model

We quantify heritable effects on drug response using GxEMM, a linear mixed model for genome-wide gene-environment interactions [1]. GxEMM quantifies the heritability contributed by genome-wide additive effects and genome-wide GxE effects. The general GxEMM model is:

$$\begin{aligned}
y &= X\alpha + G\beta + (G * Z)\gamma + (I * Z)\epsilon \\
\beta_l &\overset{\text{iid}}{\sim} \mathcal{N}\left(0, \frac{1}{L}\sigma^2_{hom}\right) \\
\gamma_{(l,)} &\overset{\text{iid}}{\sim} \mathcal{N}\left(0, \frac{1}{L}V\right) \\
\epsilon_{i,} &\overset{\text{iid}}{\sim} \mathcal{N}(0, W)
\end{aligned}$$

In this model, the known data are:

- $y$ is the quantiative phenotype
- $X$ are covariates with fixed effects $\alpha$, like age or sex.
- $G$ is a centered and scaled matrix of genome-wide genotypes
- $Z$ is matrix of context features. In our study, $Z_{i,} = (0, 1)$ if individual $i$ is treated, and $Z_{i,} = (1, 0)$ if individual $i$ is untreated
- $*$ is the column-wise Khatri-Rao product, which forms the interaction between two design matrices. For example, each column of $G * Z$ is of the form $G_{,j} \circ Z_{,k}$, where $\circ$ takes the element-wise product between SNP $j$ and context feature $k$

The random effects and their corresponding variance components are:

- $\beta_l$ is the effect of SNP $l$ that is shared across contexts. $\sigma^2_{hom}$ is the additive genetic variance—i.e., the size of $\sum_l \beta_l$
- $\gamma_{(lk)}$ is the effect of SNP $l$ that is specific to context $k$. $v_k := V_{kk}$ is the genetic variance specific to context $k$—i.e., the size of $\sum_l \gamma_{(lk)}$
- $\epsilon_{(ik)}$ is the noise for individual $i$ from context $k$. $w_k := W_{kk}$ is the noise variance in context $k$.

Finally, the cross-context covariance terms are:

- $v_{12} := V_{12}$ is the genetic covariance between contexts. Because $Z$ is binary in our setting, this term be ignored WLOG—it can be folded in with $\sigma^2_{hom}$ [1]
- $w_{12} := W_{12}$ is the noise covariance between contexts 1 and 2. In our setting with binary $Z$, this term cannot be identified—an individual either has noise $\epsilon_1$ or $\epsilon_2$, but we cannot observe the covariance between these terms in cross-sectional data.

While neither $v_{12}$ nor $w_{12}$ can be identified in our cross-sectional data, they are different in an important way. Specifically, $v_{12}$ can be assumed 0 WLOG, because it is already captured in $\sigma^2_{hom}$. However, $w_{12}$ cannot be assumed to be zero—we simply have no data to learn about this parameter.

To unpack the model, imagine studying genetic effects on LDL cholesterol across statin users and non-users. A SNP $s$ that equally increases LDL cholesterol in both groups has a homogeneous effect

($\beta_s > 0$) but has no drug-specific effects ($\gamma_{s0} = \gamma_{s1} = 0$), so $s$ contributes to $\sigma^2_{hom}$ but not $v_0$ or $v_1$. Conversely, a SNP $s'$ that increases LDL cholesterol only in statin users has $\beta_{s'} = 0$ and $\gamma_{s'1} > \gamma_{s'0} = 0$, so $s'$ contributes to $v_1$ but not $\sigma^2_{hom}$ or $v_0$. Finally, $w_1 > w_0$ means that statin users have higher non-genetic LDL cholesterol variance.

In various special cases, GxEMM is similar or identical to other methods that fit genome-wide GxE heritability [2–4]. For example, the method from [3] applies to categorical environments and continuous phenotypes and, thus, would apply to our analyses of quantitative phenotypes (such as LDL cholesterol or A1C) but would not apply to our analyses of binary disorders (such as T2D or CVD). Finally, GxEMM reduces to the ordinary additive heritability model when $V = 0$ and $w_1 = w_2$, i.e., when neither genetic nor nongenetic variance depends on the environment.

#### 3.2 Using GxEMM to estimate treatment response heritability

If we had measures of a phenotype before and after a treatment, we could directly calculate the change in phenotype, $\Delta y$, and then estimate its heritability using standard heritability estimation methods. This would be ideal, as the change in phenotype captured in $\Delta y$ cancels out the contribution of all covariates and unmodelled noise that do not depend on treatment status.

In contrast, we are interested in the setting where we only measure each individual's phenotype before or after treatment. This is motivated by large cross-sectional biobank data like UK Biobank, where most individuals are only observed at one time point. Here, we show how to approximate the heritability of $\Delta y$ in this setting using GxEMM.

How is this possible if we never observe $\Delta y$? Imagine we only see individual $i$ pre-treatment ($E_i = 0$), but that we see their relative $j$ post-treatment ($E_j = 1$). Intuitively, we can use individual $j$'s post-treatment phenotype to proxy for individual $i$'s post-treatment phenotype. More specifically, individual $j$ will be a proxy for the genetic part of individual $i$. Intuitively, individual $j$ cannot inform the nongenetic part of individual $i$'s phenotype; mathematically, this is equivalent to our above observation that $w_{12}$ is not identified.

To declutter notation, consider a single individual's phenotype $y$ and genotype vector $g$. Let $\gamma_1$ indicate the effects of all $S$ SNPs in untreated individuals ($E = 0$), and let $\gamma_2$ indicate the effects of the SNPs in treated individuals ($E = 1$). Informally define $\Delta y$ as $y(E = 1) - y(E = 0)$, i.e., the phenotype change after an individual is treated. $\Delta y$ is unobserved because we only observe either the treated or untreated state. Our goal is to estimate the heritability of this unobserved phenotype using GxEMM. Under the GxEMM model defined above, we have:

$$
\begin{aligned}
\Delta y &= y(E = 1) - y(E = 0) \\
&= (g\beta + g\gamma_2 + \epsilon_2) - (g\beta + g\gamma_1 + \epsilon_1) \\
&= g(\gamma_2 - \gamma_1) + (\epsilon_2 - \epsilon_1) \implies \\
\mathbb{V}(\Delta y) &= \operatorname{tr}\left(g^T g \mathbb{V}(\gamma_2 - \gamma_1)\right) + \mathbb{V}(\epsilon_2 - \epsilon_1) \\
&= \operatorname{tr}\left(g^T g \frac{1}{L} I_L (v_{11} + v_{22})\right) + (w_{11} + w_{22} - 2w_{12}) \\
&= (v_{11} + v_{22}) + (w_{11} + w_{22} - 2w_{12})
\end{aligned}
$$

GxEMM can estimate all of these parameters—except for $w_{12}$. Intuitively, it captures the covariance in effect sizes for unmodelled risk factors between treated/untreated states. We can safely assume that $w_{12}$ is nonnegative: Otherwise, the majority of unmodelled nongenetic risk factors would have opposite effects in the treated/untreated contexts. For example, if smoking status was the only unmodelled risk factor for LDL cholesterol, then $w_{12} < 0$ implies smoking becomes protective after statin administration.

We emphasize that this is a biological assumption, not a mathematical assumption.

Therefore, we can estimate the heritability of treatment response by:

$$\begin{aligned} h^2_{response} &:= h^2(\Delta y) \\ &= \frac{v_{11} + v_{22}}{v_{11} + v_{22} + w_{11} + w_{22} - 2w_{12}} \\ &\geq \frac{v_{11} + v_{22}}{v_{11} + v_{22} + w_{11} + w_{22}} \end{aligned}$$

We call this "conservative" in the main text to emphasize that the heritability is underestimated when $w_{12} > 0$, and in this sense the inequality is mathematically conservative. But, again, we will overestimate $h^2$ in the unlikely case where $w_{12} < 0$.

Finally, we note that these calculations ignore endogeneity in treatment status. This is hiding in our informal definition of $\Delta y$, which imagines that we observe an individual in $E = 0$ or $E = 1$ state at random. However, when the treatment is prescribed based on $y$ itself, we are ignoring a subtle form of dependence between $E$ and $G$ (that is more pernicious than mere G-E correlation, which does not generally cause bias in GxEMM [1]). This is worth theoretically solving in future work; here, we use simulations to evaluate the implications of this potential bias.

#### 3.3 Implementing GxEMM in UK Biobank

Since GxEMM can accommodate only moderate sample sizes, for warfarin, methotrexate, and metformin, we studied a sample of 30,000 individuals that included all users of that drug and an accordingly-sized random draw of non-users. To assess stability of our results, we repeated the analysis five times by randomly resampling non-users, and report results from the sample with median additive heritability ($h^2_{hom}$, Figure 4). Because statins are much more common, we instead randomly split all 342,257 individuals into 11 non-overlapping subsets and meta-analyzed the results, which is a common approach in biobank-scale datasets [5].

#### 3.4 Simulating endogenous "E"

In practice, drugs are not administered at random. When the causes of drug prescription are intertwined with causal effects on the focal phenotype, this is called endogeneity. In general, endogeneity can induce complex biases in statistical analyses. Randomized controlled trials are the gold-standard approach to solve this, as the drugs are truly administered at random, i.e., exogeneously. Developing comprehensive statistical approaches to remove endogeneity bias in gene-drug interaction studies is a valuable future direction.

Here, we perform simulations to understand a particular form of endogeneity that is surely present in our analyses: statins are prescribed for individuals with higher levels of LDL cholesterol. We are concerned with the impact of this endogeneity on our gene-drug interaction tests in the absence of any genetic interactions at baseline. Therefore, we simulate LDL cholesterol as a purely additive genetic trait, as is standard in complex trait genetics:

$$\begin{aligned} y &\sim G\beta + \epsilon \\ \beta_l &\overset{\text{iid}}{\sim} \mathcal{N}\left(0, \sigma_g^2/L\right) \\ \epsilon_l &\overset{\text{iid}}{\sim} \mathcal{N}\left(0, \sigma_e^2\right) \end{aligned}$$

We use $N = 10,000$ samples, $L = 100$ SNPs, $\sigma_g^2 = .5$, and $\sigma_e^2 = .5$. Note that this is a special case of our GxEMM model, where $V = 0$ and $W = \sigma_e^2 I_2$. That is, the genetic heterogeneity is absent, and the noise is i.i.d. across contexts.

The novel part of our simulation is adding a drug effect in a way that depends on $y$. Specifically, we assume the drug ($E$) is administered to individuals above the 80th percentile of the LDL cholesterol distribution. We consider two different models for the drug effect:

- Homogeneous across individuals. In this case, the phenotype $y$ is modified by:

  $$y \leftarrow y + E\beta_E$$

  where $E$ is a 0-1 indicator of drug use status, and we take $\beta_E = -1$. (This is one standard deviation on the pre-treatment phenotype scale.)

- Projecting individuals to the treatment threshold: all individuals with $y_i$ above the threshold are returned directly to the threshold.

These operations are visualized in the post-treatment phenotype histograms in the left column of Figure 5. These two scenarios are intended to represent two different realistic treatment effects: in the homogeneous case, everyone gets the same effect; in the threshold case, everyone is given a drug dosage/regimen to achieve a target phenotype.

After simulating the data, we then perform a series of regressions to understand the impact on effect size estimates. First, we compare the additive genetic effects (from regressing post-treatment phenotypes on $G$) to the interaction genetic effects (the interaction term from regression post-treatment phenotypes on $G \times E$). As expected based on our real data analyses in Figure 2, we find that GxE effects are starkly negatively correlated with additive effects (second column of Figure 5). This reflects systematic buffering of genetic effects after treatment, which can also be seen when we compare the effect size estimates from only treated vs only untreated individuals (third column of Figure 5).Finally, we observe that the additive effect estimates are modestly reduced when fitted to post-treatment phenotypes rather than pre-treatment phenotypes (fourth column).

We next fit GxEMM with HE regression to estimate $h^2(\Delta y)$ from the same simulations (with the Homogeneous 'E' effect for simplicity). We found that $h^2(\Delta y)$ was 13.8% under this model (on average over 100 simulations, standard error=0.6%). These simulations assume that baseline LDL cholesterol has heritability of 50%. When we instead assume baseline heritability of 20%, we found that $h^2(\Delta y)$ was 4.3% (on average over 100 simulations, standard error=0.4%). Qualitatively, these results are consistent with the observed $h^2(\Delta y)$ for the LDL cholesterol response to statins that we observe in practice (9%) because the LDL cholesterol heritability likely lies in the range of 20-50%.

### 4 TxEWAS for detection of gene-drug interactions

#### 4.1 The TxEWAS model

To identify specific genes involved in drug response from cross-sectional data, we use a newly developed statistical framework, TxEWAS, which extends transcriptome-wide association studies (TWAS [6, 7]). TxEWAS addresses shortcomings of GWAS of drug response—it improves power by reducing the number of tests, and interpretability by directly nominating causal genes.

The TxEWAS framework involves two major steps: First, gene expression levels of each gene are genetically imputed using a reference dataset (Section 4.4). Second, the interaction effect between imputed gene expression and the drug is tested.

In case where a response $y_i$ is continuous, the interaction effect ($\beta_6$) is tested in the linear regression model:

$$y_i \sim \mathcal{N}\left(\beta_0 + \sum_j \beta_{1j} c_{i,j} + \sum_j \beta_{2j} e_i c_{i,j} + \sum_j \beta_{3j} g_i c_{i,j} + \beta_4 g_i + \beta_5 e_i + \beta_6 g_i e_i, \sigma^2\right),$$

where $e_i$ and $g_i$ are drug use indicator and imputed expression of some gene for individual $i$, respectively; and $c_{i,j}$ is an element of a matrix of covariates $\boldsymbol{C}$. $\beta_4$ and $\beta_5$ are what we call "main" or "aditive" effects of a gene and drug, respectively.

In case where the response is binary, the interaction effect is tested in the logistic regression model:

$$y_i \sim \text{Bernoulli}\left(\text{logit}^{-1}\left(\beta_0 + \sum_j \beta_{1j} c_{i,j} + \sum_j \beta_{2j} e_i c_{i,j} + \sum_j \beta_{3j} g_i c_{i,j} + \beta_4 g_i + \beta_5 e_i + \beta_6 g_i e_i\right)\right).$$

The variance of the effect size estimates is estimated with the robust sandwich variance estimator (SVE). In the linear regression-based test, their role is to control for both heteroscedasticity [8] and misspecification of the functional form of the environmental factor [9, 10]. In the logistic regression-based test, they are meant to account for the latter. The bias caused by the violation of the homoscedasticity assumption of the linear regression model is a specifically important concern in GxE studies. We therefore perform extensive simulations and permutation analyses to test that the TxEWAS model is calibrated (Section 4.2).

We note that even though in this work we use TxEWAS to detect gene-drug interactions, any environment can be tested with this method.

#### 4.2 TxEWAS performance in simulations

In the presence of environment-conditional heteroscedasticity, testing multiple genetic variants for interaction with the environmental factor in a simple linear regression model results in an inflated or deflated false positive rate (FPR), depending on the relation between group size and phenotypic variation [8]. We performed a simulation to assess the size of this bias in a dataset of the size and characteristics of the UK Biobank (Figure 1A).

In the simulation, we considered a binary environmental variable that divided observations into two groups of sizes $n_1$ and $n_2$, and phenotype variances $\sigma_1^2$ and $\sigma_2^2$. We simulated the phenotype as a Gaussian random variable with the corresponding group variances, but with no mean effects. The genotype was drawn from a binomial distribution with the same probability 0.4 of success for both groups. We fitted to this data GxE models that included the genotype, the environmental factor and the product of those two as covariates. For every selected value of the ratio $\sigma_2^2/\sigma_1^2$, we ran 5,000 such simulations, and calculated the FPR for the GxE effect as the proportion of simulations where the nominal P value for this effect was less than 0.05.

Group sizes used in the simulation were chosen based on the number of statin users in the UK Biobank ($n_1 = 285,822$ and $n_2 = 56,132$). Examples of $\sigma_2^2/\sigma_1^2$ values that we encountered in the UK Biobank when stratifying individuals by statin use were: 0.68 for LDL cholesterol, 1.96 for blood glucose, or 3.00 for A1c.

The simulation shows that if the smaller (larger) group is characterized by the larger (smaller) variance of the response, the FPR for the GxE model fitted with ordinary least-squares (OLS) is inflated (deflated). Within a realistic range of parameter values, the FPR can reach zero or increase threefold (Figure 1A). On the other hand, if the groups have equal sizes, the model is well calibrated. The bias can be controlled by using robust standard errors estimated with the sandwich variance estimator (SVE). This approach gives virtually the same results as the double generalized linear model (DGLM), which correctly models the phenotypic variance as a function of covariates, and is more computationally efficient than the latter.

Taking advantage of those facts, TxEWAS performs a linear regression-based interaction test and controls for heteroskedasticity with the SVE.

To investigate the type I error rate for TxEWAS in real data, we performed a permutation analysis where we randomly shuffled imputed expression of 4,516 liver genes across subjects. We selected statin for drug exposure, and LDL cholesterol and blood glucose for responses to examine both deflation and inflation biases ($\sigma_2^2/\sigma_1^2 = 0.68$ and $\sigma_2^2/\sigma_1^2 = 1.96$, respectively). The analysis demonstrates that the TxEWAS model is well calibrated, even though P values estimated with the simple linear regression-based interaction test are heavily deflated or inflated, depending on the examined response (Figure 1B).

#### 4.3 Performing TxEWAS in UK Biobank

For this study, we imputed gene expression into the UK Biobank using the 48 tissues from the GTEx consortium [11], and tested genes whose expression was significantly predicted at a nominal P value of 0.05 (Section 4.4). To call significant interactions at FDR $< 10\%$, we applied the hierarchical FDR (hFDR) correction with the treeQTL software [12, 13]. hFDR has been shown to properly control the false discovery rate across contexts when there are multiple hypothesis tests being run for a given group, in this case, a gene [12–14]. It also boosts power in cases where a gene has a significant association in multiple contexts. While this careful approach to combining tissues is important for the sake of calibrated statistical testing, when we visualize effect sizes we simply use the tissue with the most significant interaction effect.

Finally, since TxEWAS is liable to detect genes that are merely correlated with the causal gene due to the genetic LD structure in the proximity of the causal gene, we define TxEWAS association loci by adding consecutive genes until there is no gene within 500 kb from the last added gene.

#### 4.4 Gene expression prediction models

In TxEWAS, gene expression (or rather its genetic component) is imputed as a linear combination of genetic variants (SNPs). The coefficients used for the imputation are referred to as "weights" or "eQTL weights" and are calculated on a per-gene basis by fitting a linear model of gene expression onto the gene's *cis*-genotypes in an external reference dataset. We fit each model using the elastic net, as it has been found to be the most robust across a wide range of genetic architectures [15]. We used package bigstatsr [16] to fit each model using 10-fold cross-validation, and after determining whether the expression of a gene was significantly predicted using *cis*-genotypes at a nominal P value of 0.05, we retrained the model using the entire set of individuals to generate a final set of weights.

#### 4.5 Replication in a pharmacogenomic study

To replicate our UK Biobank findings we evaluated TxEWAS associations in a traditional pharmacogenomic study. The study assessed genome-wide effects of genetic variants on statin-induced LDL cholesterol change. The phenotype was rigorously characterized utilizing electronic health records in a multiethnic population of 34,874 statin users from the Kaiser Permanente GERA cohort (Genetic Epidemiology Research on Adult Health and Aging), and the analysis was adjusted for carefully selected covariates [17, 18]. We used summary statistics for European individuals ($n = 28,616$) from this study to perform a transcriptome-wide association study (TWAS) [6, 7] for genes identified in the statin-LDL TxEWAS. This TWAS tested the *cis* genetic component of expression of these genes for association with variability of LDL cholesterol reduction rate after statin initiation. For every gene, we calculated the TWAS statistic in all available GTEx tissues, and employed an hFDR correction ($\alpha < 10\%$) to call statistically significant genes.

We performed this TWAS using FUSION [7] and the LD reference data for individuals of European ancestry provided with the software. The LD reference was calculated based on genotypes from the 1000 Genomes Project, which provided fewer SNPs than the UK Biobank. As a result, for some of the genes that we identified with TxEWAS using the individual-level UK Biobank data, the expression models could not be built by FUSION for the pharmacogenomic study. This is why, in the main analysis, we report replication rate relative to 155 genes, and not all 156 genes identified in the statin-LDL TxEWAS.

##### 4.6 Combinations of treatments

Patients are often on multiple drugs simultaneously, for example, in the cohort studied in this work, out of the 8,606 individuals who reported taking metformin 6,946 also reported taking statins. This makes accompanying treatments an important potential confounder in our analyses. To test their impact on GxE, we repeated TxEWAS for each drug excluding individuals on the other drugs we consider—e.g., in the TxEWAS for statins, we excluded individuals taking metformin, warfarin, or methotrexate. We observe that the original results remain largely unaffected (Figure S4). For example, the statins-LDL analysis identified 127 interaction genes, from which 113 were among the 156 interaction genes found in the original analysis. Of those 127 genes, 126 could be studied in the replication cohort, and 35 replicated at hFDR < 10%. The remaining genes were enriched for low P values < 0.1 (binomial test=0.002).

#### 5 The impact of gene-drug interactions on polygenic prediction accuracy

##### 5.1 Assessing implications for polygenic scores in clinical practice

We assessed transferability of PGS between samples that are of similar genetic ancestry but differ by the drug use status. We considered four settings for the training data: 1) users only, 2) non-users only, 3) all users and all non-users, and 4) half of users and half of non-users (randomly subsampled to match sizes of sets 1 and 2). We trained two PGS in individuals from set 4: "standard-PGS", where we used the standard covariates (age, sex, birth date, Townsend deprivation index, and the first 16 genetic PCs); and "half-PGS", where we added an indicator of drug use to the standard covariates. The drug use indicator was also added when training PGS in all users and all non-users ("all-PGS"). We fitted PGS using a fast implementation of penalized linear regression with the lasso penalty [19, 20], and we measured prediction accuracy by the incremental $R^2$ over baseline covariates (Table 5). Standard errors around the estimates were calculated using bootstrap.

##### 5.2 Simulating polygenic scores

We performed realistic simulations to examine two scenarios observed in real data. Scenario 1 mimicked statins-LDL, where "on" genetic effects are buffered to be half the size of "off" effects. As expected, we found that training PGS in "off" individuals is optimal regardless of the test set (Table 6). Scenario 2 mimicked statins-A1c, where "on" and "off" effects are highly correlated but vary randomly in magnitude. As expected, we found that training PGS in training samples matching the test samples is optimal in this scenario. These simulations show how prediction accuracy of a PGS depends on the genetic correlation and heritability between the train and test dataset and explain the discordant results for statins' effects on A1c and LDL cholesterol (Table 5).

In both cases, we simulated the unexposed population to have 40% heritability. In the exposed population, we either (Scenario 1) divided all genetic effect sizes by two, reflecting systemic buffering of the unexposed effects, or (Scenario 2) randomly deflated (with probability 0.4) or inflated (with probability 0.6) each individual genetic effect by a random fraction between 0.2 and 1. We then performed PRS analyses as in the real data by varying the distribution of drug use in the train and test populations.

### 6 Data

#### 6.1 Samples

Analyses presented in this work were performed in the UK Biobank population of 342,257 unrelated white British individuals, identified by performing the following steps.

From among 488,363 UK Biobank participants, we retained putative "White British" individuals using field 22006-0.0 ($n = 409,692$). We then filtered out 199 individuals with excess genotype missingness ($> 0.05$), 312 individuals with a mismatch between self-reported and genetic sex, 999 individuals with excess heterozygosity ($\geq 5$ standard deviations above the mean), and 90 individuals who requested their data be redacted. We then removed 629 individuals related to ten or more individuals (KING coefficient $\geq 2^{-9/2}$) as a preprocessing step to the application of the `maximal_independent_set` algorithm implemented in the NetworkX Python package [21].

In contrast to Bycroft et al. [22], who estimated kinships using approximately 92,000 common SNPs with small loadings onto the first few principal components (PCs) in the full sample (including multiple ancestries; see S3.7 of Bycroft et al.), we estimated kinships using 561,780 common SNPs in a sample of European ancestry individuals. The close relatives the UK Biobank identified in field 22021-0.0 are a subset of our more conservative approach: we identified all 81,218 related individuals in this subsample identified by the UK Biobank plus an additional 3,261 not identified by Bycroft et al.

#### 6.2 Genotypes

For heritability and PGS analyses, we used 579,566 UK Biobank variants with minor allele frequency (MAF) larger than 0.01, Hardy-Weinberg equilibrium (HWE) test P value below $10^{-10}$, and imputation INFO score of 1.

For the TxEWAS analysis, UK Biobank SNPs that matched eQTLs trained in the GTEx consortium [11] were used.

#### 6.3 Phenotypes

Individuals who take statins were identified by UK Biobank field 20003-0.0-47 using the following codes: 1140861958, 1140861970, 1141146138, 1140888594, 1140888648, 1140910632, 1140910654, 1141146234, 1141192410, 1141192414, 1141188146, 1140881748 and 1140864592. There were 56,169 such subjects within the UK Biobank population of 342,257 unrelated white British individuals. Individuals who take metformin ($n = 8,606$) were identified by codes 1140884600 and 1141189090 in the same UK Biobank field. Warfarin users ($n = 3,753$) were identified by codes 1140888266 and 1140910832; and methotrexate users ($n = 1,865$) by codes 1140869848 and 1140910036.

To retrieve corrected LDL cholesterol levels, we used UK Biobank fields 30780-0.0 and 30783-0.0. To obtain corrected glucose levels, we used fields 30740-0.0 and 30743-0.0. A1c, BMI, hematocrit, plateletcrit, reticulocyte count and C-reactive protein levels were retrieved from UK Biobank fields 30750-0.0, 21001-0.0, 30030-0.0, 30090-0.0, 30240-0.0 and 30710-0.0, respectively.

C-reactive protein levels were inverse normally transformed before fitting the models. For other traits, we discarded measurements greater than five standard deviations from the mean, with the assumption that such extreme levels were results of non-modeled circumstances.

CVD was defined as in Thompson et al. [23]. The T2D disease status was extracted from the UK Biobank electronic health records using E11.0-E11.9 ICD10 codes. For testing associations with drug use, we only retained diagnoses recorded after the date of the initial assessment with the UK Biobank initiative (when the information about medication use was collected).

#### 6.4 Covariates

The main analyses reported in this work were performed using the following covariates: age, sex, birth date, Townsend deprivation index, and the first 16 genetic PCs [20]. We additionally accounted for the measuring device type when an outcome required it, which was the case for hematocrit, plateletcrit, and reticulocyte count. To simplify interpretation, all non-binary covariates were standardized (transformed to mean-zero, variance 1) before calculating interaction variables.
